# Social Distancing Has Merely Stabilized COVID-19 in the US

**DOI:** 10.1101/2020.04.27.20081836

**Authors:** Aaron B. Wagner, Elaine L. Hill, Sean E. Ryan, Ziteng Sun, Grace Deng, Sourbh Bhadane, Victor Hernandez Martinez, Peter Wu, Dongmei Li, Ajay Anand, Jayadev Acharya, David S. Matteson

## Abstract

Social distancing measures, with varying degrees of restriction, have been imposed around the world in order to stem the spread of COVID-19. In this work we analyze the effect of current social distancing measures in the United States. We quantify the reduction in doubling rate, by state, that is associated with social distancing. We find that social distancing is associated with a statistically-significant reduction in the doubling rate for all but three states. At the same time, we do not find significant evidence that social distancing has resulted in a reduction in the number of daily confirmed cases. Instead, social distancing has merely stabilized the spread of the disease. We provide an illustration of our findings for each state, including point estimates of the effective reproduction number, *R*, both with and without social distancing. We also discuss the policy implications of our findings.

## 1 Introduction

Over nine hundred thousand Americans are confirmed to have contracted SARS-CoV-2, the virus that causes COVID-19, as of April 25th, 2020 [1]. Efforts to stem the spread of the disease have resulted in unprecedented societal disruptions [2, 3, 4, 5, 6, 7, 8, 9]. These efforts have largely taken the form of mandated or recommended social distancing measures by state and local governments. These social distancing measures incur substantial economic cost [10, 11]. At the same time, their efficacy in reducing the spread of the disease is presumed but not well understood. Quantifying the benefits of social distancing measures is of paramount importance as policy makers consider how and when to relax such measures.

Around mid-March, the fifty states and the District of Columbia independently imposed a similar suite of social distancing measures at nearly the same time. These measures appear to have caused a substantial, albeit delayed, change to the spread of the virus (See Fig. 1). By separately analyzing the spread of the virus before and after these interventions, appropriately correcting for delays in their effect, one can quantify and test their effectiveness.

**Figure 1:**
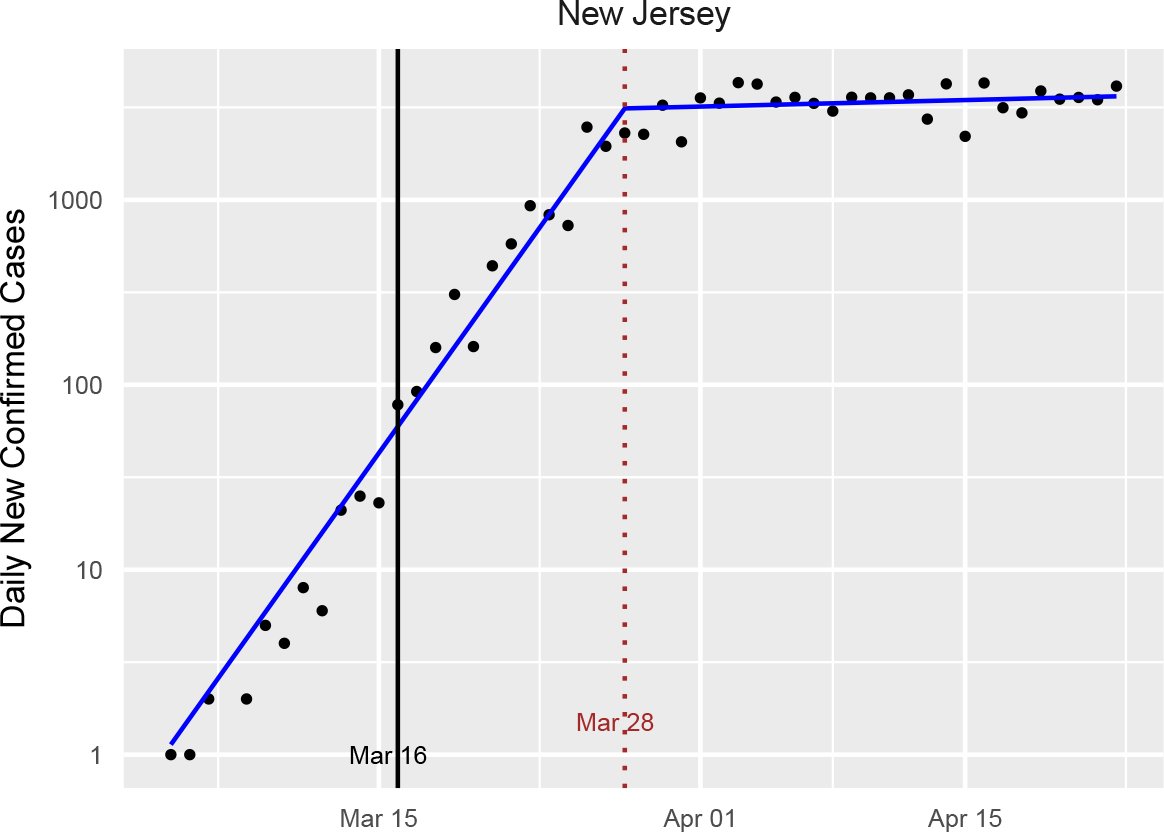
New confirmed cases of COVID-19 by day for New Jersey, along with the fitted model from our *learned-delay* analysis. Under our definition, New Jersey began social distancing on March 16th, and a clear changepoint is observed around 12 days later. A 12-day delay between infection and confirmed test is consistent with other states and with the independent estimate in Appendix A. Note that the number of new cases per day appears to have merely plateaued as a result of the intervention. We call this the *standard plot* for New Jersey. Standard plots for all states and the District of Columbia are available in Appendix B.

### 1.1 Contributions and Findings

Using an interrupted time-series analysis of the confirmed case counts, we estimate the doubling rate of the spread across the different states in the US both before and after the imposition of widespread social distancing measures (hereafter, the “intervention”). Compensating for the delay between infection and confirmed diagnosis is a central component of the analysis. We find that in all states except Nebraska, North Dakota, and South Dakota, the intervention is associated with a statistically-significant decrease in the doubling rate of the spread. The average of the pre-intervention doubling-rate point estimates across the states was 0.302 days^*−*1^, meaning that the number of new infections per day was doubling every 3.31 days. Post-intervention, the average fell to 0.010 days^*−*1^, meaning that the number of infections was still increasing, but required about 100 days to double. We are not able to conclude that any of the states made the spread subcritical (i.e., obtained a negative doubling rate) through the interventions, although some have negative point estimates of the post-intervention doubling rate. Thus while this study finds substantial evidence that social distancing interventions reduce the reproduction rate of SARS-CoV-2, it also suggests that these measures are insufficient to make the spread subcritical. That is, social distancing has largely plateaued, but not decreased, the number of infected individuals. The policy implications of this finding are discussed in Section 5.

As part of the analysis we estimate the average delay between infection and confirmed diagnosis using two different methods. These give estimates of 11.5 and 12 days, respectively, which are consistent with an estimate obtained by combining estimates from the literature on the length of the incubation period and the time between symptom onset and positive test, as described in Appendix A.

### 1.2 Related Work

Estimating the doubling rate of a disease like COVID-19 is related to the problem of estimating the mean number of “descendants” in a Galton-Walton branching process [12]. Classical methods [13, 14] for that problem assume a time-invariant distribution over the number of descendants, which differs from the change-point method used here. Wallinga and Teunis [15] provide an approach for estimating the number of people infected by individual patients with an infectious disease, not merely the average over this group. By considering when specific patients were known to be infected, they obtain an estimate of the infection rate over time. They assume a closed system, however, in which there are no external infections beyond the first. Bettencourt and Ribeiro [16] note that this method requires a large number of samples, and they propose a Bayesian alternative requiring less data. A recent extension of their approach has been applied to COVID-19 tracking in the US [17]. Their method (and the dashboard in [17]) does not account for interventions, so it does not provide separate pre- and post-intervention estimates of parameters, which are necessary for determining the efficacy of the intervention. Intervention-aware methods, such as those considered here, also provide more accurate real-time estimates by separating data that comes from different regimes. The concern that the Bettencourt and Ribeiro estimator would be slow to reflect interventions has already been raised [17].

The closest work to the present paper is that of Siedner *et al*. [18], which also examines the spread of COVID-19 in the US using an interrupted time series analysis with the goal of quantifying the effect of social distancing. They find the impact of social distancing to be relatively modest, increasing the doubling time from 3.3 days to 5.0 days. They implicitly assume the average time between infection and confirmed test, which we call *confirmed case delay*, to be 4 days. They also rely on data only up to (and including) March 30th. Our estimates place the confirmed case delay at around 12 days, so that the effects of the interventions would not appear until late March. By allowing for a 12 day delay and using data up to and including April 23rd, we conclude that social distancing interventions are associated with a much larger reduction in the spread of the disease than that found by Siedner *et al*.

Interrupted time series (ITS) analysis, which is the primary statistical method used in this work, is a quasi-experimental study design that has been increasingly used to evaluate the effectiveness of policy changes on longitudinal data in public health [19]. In particular, ITS analysis has been used to evaluate health care quality improvement using population-level longitudinal data [20]. Existing work has also used ITS analysis to examine the effectiveness of the transition from ICD-9-CM to ICD-10-CM coding on injury hospitalization [21].

## 2 Data

We have used multiple independent data sources for the various analyses reported in the paper. State-level case and death counts were obtained from the New York Times (NYT) Github repository [1]. This data includes only lab-confirmed cases of COVID-19 reported by a federal, state, territorial, or local government agency. Thus it does not count cases that are probable but not confirmed. As only a small set of states are reporting probable cases (e.g., Ohio, Wyoming, and Idaho) starting after April 14, 2020, we rely on confirmed cases in order to be consistent across states.

We have used K-12 school [22] and restaurant [23] closures as indicators of interventions. These were chosen because they represent the first widely-disruptive social distancing measures that were imposed. If a school closure was announced in the evening, we consider it as applying the next day. If closure was announced on a weekend, we used the first weekday for which schools were closed. The earlier of restaurant closing and school closing is defined as the *intervention date* for each state (Table 4). Taking the earlier of the two reduces the effect of anticipatory behavior.

For the analysis in Appendix A on the confirmed case delay, we use data on COVID-19 cases by illness onset recorded by the CDC [24]. We reference data on testing in New York State in Section 6. This data was obtained from the COVID Tracking Project [25].

## 3 Methodology

We focus on estimating the doubling rate, *β*, measured in inverse days, of new infections across the fifty states and the District of Columbia^1^, both before and after the intervention. We assume that during each period, the number of new infections per day is expressed as

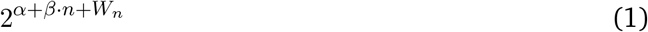

where *n* is a discrete time index, *α* is a constant, and {*W*_*n*_} is a mean-zero Gaussian noise process. The parameters *α* and *β* are assumed to vary both with the intervention and across states. Note that *β* can be negative, which corresponds to a contraction of the number of cases with time. Adopting the terminology of branching processes [12], we call the process *supercritical* if *β >* 0, *critical* if *β* = 0, and *subcritical* if *β <* 0.

When analyzing pandemic spreads, it is common to infer the *effective reproduction number, R*, instead of the doubling rate. We focus on the doubling rate because it can be more directly estimated from the time-series of confirmed cases. One can compute *R* from the doubling rate *β* and the distribution of the serial interval using the Lotka–Euler equation [26, 27]. Applying Jensen’s inequality to this equation gives an upper bound on *R* in terms of *β* and the mean serial interval *µ* [28],

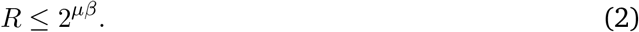

Prior work has estimated *µ* at 4 days for COVID-19 [29, 30]. All of the doubling-rate estimates in this paper can be translated to estimated upper bounds on *R* using this inequality, although the resulting bounds are less certain than the underlying estimates of *β* due to the exponentiation and uncertainty about *µ*.^2^

Considering each state in isolation, we estimate the doubling rate before and after the intervention as follows. Let *C*_*n*_ denote the number of new confirmed cases on day *n*, where the index *n* = 1 refers to the first day after March 1st for which the state records at least two consecutive days of positive confirmed cases. This convention is designed to appropriately handle states with one very early case (e.g., Washington) followed by many days of zero cases. We apply the transformation

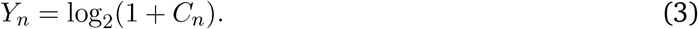

Including the effect of the intervention, we model *{Y*_*n*_*}* as

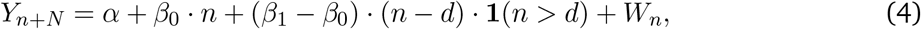

where **1**(*·*) is the standard indicator function, and *{W*_*n*_*}* satisfies

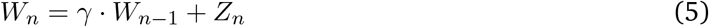

where {*Z*_*n*_} is an independent and identically distributed (i.i.d.) Gaussian sequence with mean zero and variance *σ*^2^, making *{W*_*n*_*}* a first-order autoregressive [AR(1)] Gaussian process. The parameter *d* represents the state-specific intervention date defined in the previous section. Thus *d* is known but state-dependent. The quantity *N*, which we call the *confirmed case delay*, represents the average time (in days) between when someone is infected with SARS-CoV-2 and when they are confirmed to have COVID-19 and included in the published counts. We assume that *N* is integer-valued. The parameters *β*_0_ and *β*_1_ are the doubling rates before and after, respectively, the intervention. The unknown parameters in the model are

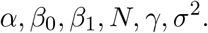

We are particularly interested in *β*_0_, *β*_1_, and *N*.

For each of the states, we fit the model in two ways, which differ primarily in how they handle the confirmed case delay, *N*. The first, which we call the *learned-delay* approach, fits *N* to the data on par with the other parameters in the model. This is tantamount to finding a change-point in the {*Y*_*n*_} time series for which there is a significant difference between the doubling rates. The difference between the changepoint epoch and *d* then forms a point estimate of *N*. We use the algorithm in [31] for this purpose, which declines to identify a changepoint (effectively setting *β*_1_ = *β*_0_) if there is insufficient evidence of a change. For states with a detected changepoint, we then obtain a point estimate of *N* in addition to the other parameters, which can vary by state. This approach assumes *γ* = 0, so that the errors are taken to be i.i.d.

The second approach, which we call *fixed-delay*, takes the confirmed case delay to be *N* = 12 for all states. This choice can be justified as follows. First, 11.5 is the median estimate of *N* across the states obtained by the learned-delay method (See Fig. 3). Second, estimating the confirmed case delay from a separate analysis of CDC data gives a point estimate 12 days, as described in Appendix A. The fixed-delay method does not require that *γ* = 0, i.e., it allows for *{W*_*n*_*}* to be AR(1). For the fixed-delay method, we conducted a sensitivity analysis on the order of the autoregressive noise process and found that allowing for higher-order dependence did not significantly improve the fit. For both methods, the unknown parameters are estimated via maximum likelihood.

**Figure 2:**
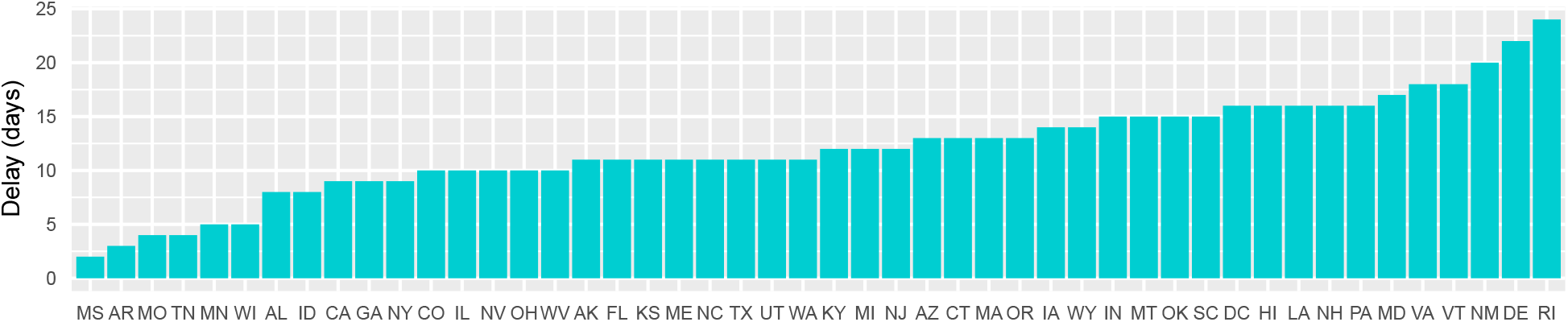
Time between the defined intervention and identified changepoint. This forms an estimate of the confirmed case delay *N* for each state. Three states (Nebraska, North Dakota, and South Dakota) do not have significant changepoints and are not included.

**Figure 3:**
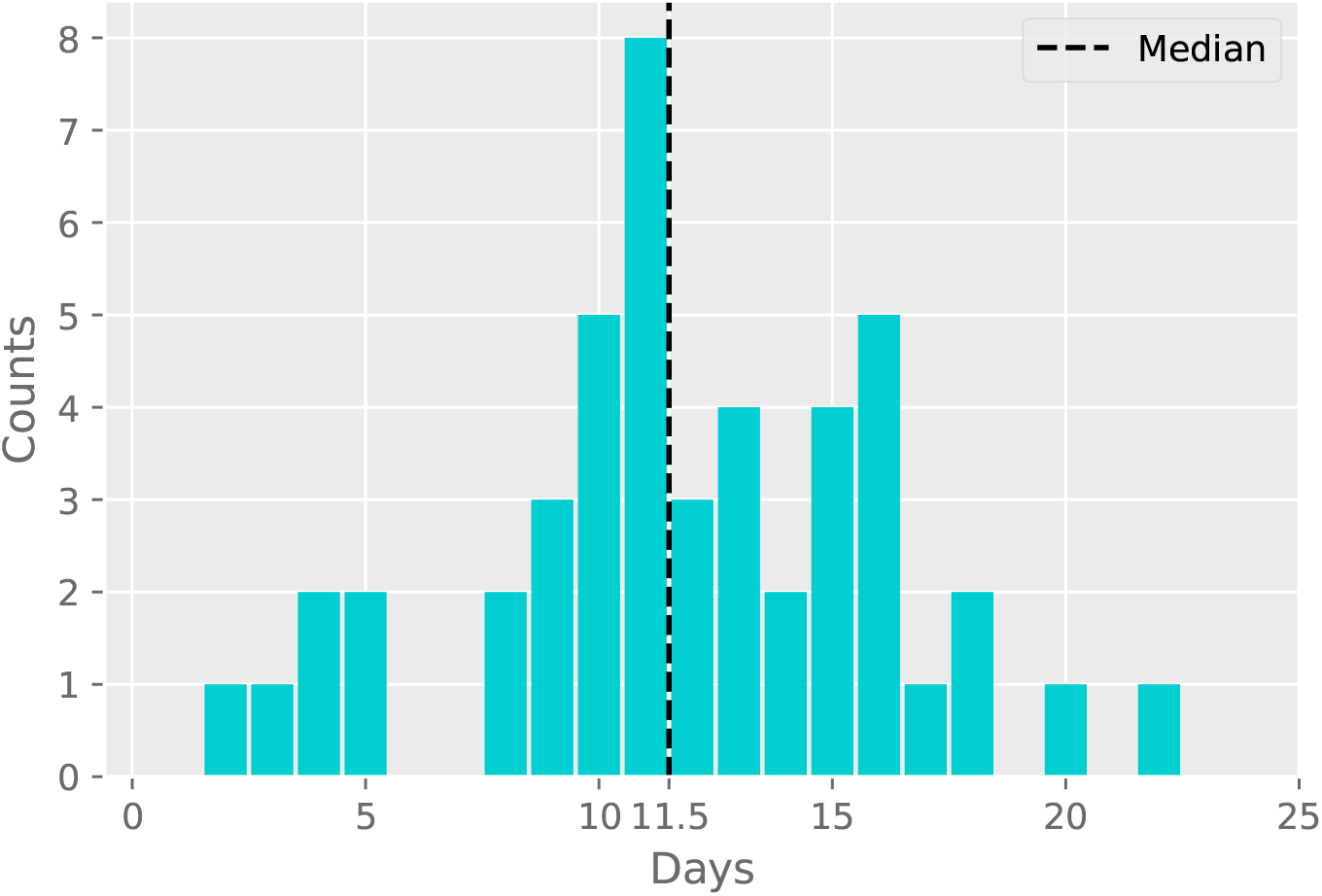
Histogram of point estimates of the confirmed case delay *N* given in Fig. 2.

## 4 Results

Tables 1 and 2 in Appendix B show the results of the learned-delay model and fixed-delay model, respectively. Fig. 6 compiles all of the findings from both models.

**Table 1:** Results from the Learned-Delay model.

| State | Intervention Date | Change Date | $N$ | $\beta_0$ | $\beta_1$ | $\beta_1 - \beta_0$ |
| --- | --- | --- | --- | --- | --- | --- |
| Alabama | 2020-03-18 | 2020-03-26 | 8 | 0.338 | 0.047 | -0.291 |
| Alaska | 2020-03-16 | 2020-03-27 | 11 | 0.221 | -0.045 | -0.266 |
| Arizona | 2020-03-16 | 2020-03-29 | 13 | 0.314 | 0.014 | -0.301 |
| Arkansas | 2020-03-17 | 2020-03-20 | 3 | 0.452 | 0.045 | -0.407 |
| California | 2020-03-19 | 2020-03-28 | 9 | 0.288 | 0.031 | -0.257 |
| Colorado | 2020-03-17 | 2020-03-27 | 10 | 0.296 | 0.008 | -0.287 |
| Connecticut | 2020-03-16 | 2020-03-29 | 13 | 0.4 | 0.041 | -0.359 |
| Delaware | 2020-03-16 | 2020-04-07 | 22 | 0.208 | 0.038 | -0.169 |
| D.C. | 2020-03-16 | 2020-04-01 | 16 | 0.258 | 0.028 | -0.23 |
| Florida | 2020-03-17 | 2020-03-28 | 11 | 0.395 | -0.015 | -0.41 |
| Georgia | 2020-03-18 | 2020-03-27 | 9 | 0.351 | 0.037 | -0.313 |
| Hawaii | 2020-03-16 | 2020-04-01 | 16 | 0.182 | -0.12 | -0.302 |
| Idaho | 2020-03-23 | 2020-03-31 | 8 | 0.295 | -0.112 | -0.407 |
| Illinois | 2020-03-16 | 2020-03-26 | 10 | 0.426 | 0.052 | -0.374 |
| Indiana | 2020-03-16 | 2020-03-31 | 15 | 0.37 | 0.0 | -0.37 |
| Iowa | 2020-03-17 | 2020-03-31 | 14 | 0.217 | 0.088 | -0.129 |
| Kansas | 2020-03-17 | 2020-03-28 | 11 | 0.285 | 0.052 | -0.233 |
| Kentucky | 2020-03-16 | 2020-03-28 | 12 | 0.24 | 0.063 | -0.177 |
| Louisiana | 2020-03-16 | 2020-04-01 | 16 | 0.358 | -0.104 | -0.462 |
| Maine | 2020-03-18 | 2020-03-29 | 11 | 0.204 | -0.031 | -0.234 |
| Maryland | 2020-03-16 | 2020-04-02 | 17 | 0.304 | 0.035 | -0.269 |
| Massachusetts | 2020-03-17 | 2020-03-30 | 13 | 0.333 | 0.047 | -0.286 |
| Michigan | 2020-03-16 | 2020-03-28 | 12 | 0.504 | -0.027 | -0.531 |
| Minnesota | 2020-03-17 | 2020-03-22 | 5 | 0.305 | 0.062 | -0.242 |
| Mississippi | 2020-03-20 | 2020-03-22 | 2 | 0.615 | 0.06 | -0.555 |
| Missouri | 2020-03-23 | 2020-03-27 | 4 | 0.466 | 0.002 | -0.464 |
| Montana | 2020-03-16 | 2020-03-31 | 15 | 0.181 | -0.121 | -0.302 |
| Nebraska | 2020-04-03 | N/A | N/A | 0.147 | 0.147 | N/A |
| Nevada | 2020-03-16 | 2020-03-26 | 10 | 0.342 | 0.0 | -0.342 |
| New Hampshire | 2020-03-16 | 2020-04-01 | 16 | 0.177 | 0.018 | -0.159 |
| New Jersey | 2020-03-16 | 2020-03-28 | 12 | 0.476 | 0.008 | -0.468 |
| New Mexico | 2020-03-16 | 2020-04-05 | 20 | 0.189 | 0.03 | -0.159 |
| New York | 2020-03-16 | 2020-03-25 | 9 | 0.5 | -0.015 | -0.515 |
| North Carolina | 2020-03-16 | 2020-03-27 | 11 | 0.371 | 0.032 | -0.34 |
| North Dakota | 2020-03-16 | N/A | N/A | 0.086 | 0.086 | N/A |
| Ohio | 2020-03-15 | 2020-03-25 | 10 | 0.435 | 0.068 | -0.367 |
| Oklahoma | 2020-03-17 | 2020-04-01 | 15 | 0.301 | -0.031 | -0.332 |
| Oregon | 2020-03-16 | 2020-03-29 | 13 | 0.204 | -0.017 | -0.221 |
| Pennsylvania | 2020-03-16 | 2020-04-01 | 16 | 0.37 | -0.001 | -0.371 |
| Rhode Island | 2020-03-16 | 2020-04-09 | 24 | 0.235 | 0.031 | -0.204 |
| South Carolina | 2020-03-16 | 2020-03-31 | 15 | 0.291 | -0.029 | -0.32 |
| South Dakota | 2020-03-16 | N/A | N/A | 0.153 | 0.153 | N/A |
| Tennessee | 2020-03-20 | 2020-03-24 | 4 | 0.415 | 0.024 | -0.391 |
| Texas | 2020-03-21 | 2020-04-01 | 11 | 0.316 | -0.003 | -0.318 |
| Utah | 2020-03-16 | 2020-03-27 | 11 | 0.303 | 0.011 | -0.292 |
| Vermont | 2020-03-17 | 2020-04-04 | 18 | 0.188 | -0.202 | -0.39 |
| Virginia | 2020-03-16 | 2020-04-03 | 18 | 0.258 | 0.058 | -0.2 |
| Washington | 2020-03-16 | 2020-03-27 | 11 | 0.214 | -0.071 | -0.285 |
| West Virginia | 2020-03-16 | 2020-03-26 | 10 | 0.423 | 0.017 | -0.406 |
| Wisconsin | 2020-03-17 | 2020-03-22 | 5 | 0.449 | 0.025 | -0.424 |
| Wyoming | 2020-03-19 | 2020-04-02 | 14 | 0.141 | -0.082 | -0.224 |

**Table 2:** Results from the Fixed-Delay model.

| State | $\beta_0$ | 95% CI | $\beta_1$ | 95% CI | $\beta_1 - \beta_0$ | 95% CI |
| --- | --- | --- | --- | --- | --- | --- |
| Alabama | 0.268 | (0.22, 0.32) | 0.027 | (-0.09, 0.15) | -0.241*** | (-0.31, -0.17) |
| Alaska | 0.196 | (0.14, 0.25) | -0.048 | (-0.18, 0.08) | -0.243*** | (-0.32, -0.17) |
| Arizona | 0.353 | (0.3, 0.4) | 0.021 | (-0.11, 0.15) | -0.332*** | (-0.41, -0.25) |
| Arkansas | 0.229 | (0.15, 0.31) | 0.021 | (-0.17, 0.22) | -0.208*** | (-0.33, -0.09) |
| California | 0.26 | (0.25, 0.27) | 0.0003 | (-0.04, 0.04) | -0.26*** | (-0.29, -0.23) |
| Colorado | 0.304 | (0.27, 0.34) | -0.012 | (-0.11, 0.09) | -0.316*** | (-0.38, -0.25) |
| Connecticut | 0.423 | (0.39, 0.45) | 0.05 | (-0.02, 0.12) | -0.373*** | (-0.42, -0.33) |
| Delaware | 0.297 | (0.21, 0.38) | 0.065 | (-0.14, 0.27) | -0.232*** | (-0.36, -0.11) |
| D.C. | 0.308 | (0.27, 0.35) | 0.055 | (-0.04, 0.15) | -0.253*** | (-0.31, -0.2) |
| Florida | 0.396 | (0.37, 0.43) | -0.025 | (-0.11, 0.06) | -0.42*** | (-0.47, -0.37) |
| Georgia | 0.333 | (0.29, 0.37) | 0.011 | (-0.09, 0.12) | -0.321*** | (-0.39, -0.25) |
| Hawaii | 0.242 | (0.17, 0.31) | -0.086 | (-0.25, 0.07) | -0.328*** | (-0.42, -0.23) |
| Idaho | 0.251 | (0.17, 0.33) | -0.152 | (-0.38, 0.08) | -0.402*** | (-0.56, -0.25) |
| Illinois | 0.403 | (0.37, 0.43) | 0.034 | (-0.04, 0.11) | -0.369*** | (-0.42, -0.32) |
| Indiana | 0.404 | (0.35, 0.46) | 0.037 | (-0.11, 0.18) | -0.367*** | (-0.46, -0.28) |
| Iowa | 0.232 | (0.18, 0.28) | 0.097 | (-0.03, 0.22) | -0.134*** | (-0.21, -0.05) |
| Kansas | 0.269 | (0.22, 0.31) | 0.05 | (-0.06, 0.16) | -0.219*** | (-0.29, -0.15) |
| Kentucky | 0.274 | (0.24, 0.31) | 0.059 | (-0.04, 0.15) | -0.215*** | (-0.27, -0.16) |
| Louisiana | 0.425 | (0.37, 0.48) | -0.047 | (-0.19, 0.1) | -0.472*** | (-0.56, -0.38) |
| Maine | 0.193 | (0.14, 0.24) | -0.035 | (-0.16, 0.09) | -0.227*** | (-0.31, -0.15) |
| Maryland | 0.361 | (0.33, 0.4) | 0.077 | (-0.01, 0.17) | -0.284*** | (-0.34, -0.23) |
| Massachusetts | 0.342 | (0.31, 0.38) | 0.056 | (-0.04, 0.15) | -0.286*** | (-0.35, -0.22) |
| Michigan | 0.494 | (0.41, 0.58) | -0.023 | (-0.23, 0.19) | -0.518*** | (-0.64, -0.39) |
| Minnesota | 0.206 | (0.17, 0.25) | 0.044 | (-0.06, 0.14) | -0.162*** | (-0.22, -0.1) |
| Mississippi | 0.248 | (0.18, 0.31) | 0.023 | (-0.14, 0.18) | -0.225*** | (-0.32, -0.13) |
| Missouri | 0.314 | (0.25, 0.38) | -0.075 | (-0.26, 0.11) | -0.389*** | (-0.52, -0.26) |
| Montana | 0.279 | (0.21, 0.35) | -0.109 | (-0.27, 0.05) | -0.388*** | (-0.48, -0.3) |
| Nebraska | 0.152 | (0.14, 0.17) | 0.092 | (-0.01, 0.2) | -0.06 | (-0.16, 0.04) |
| Nevada | 0.309 | (0.26, 0.36) | -0.002 | (-0.13, 0.13) | -0.311*** | (-0.39, -0.23) |
| New Hampshire | 0.22 | (0.19, 0.25) | 0.04 | (-0.05, 0.13) | -0.18*** | (-0.24, -0.12) |
| New Jersey | 0.485 | (0.46, 0.51) | 0.006 | (-0.05, 0.06) | -0.478*** | (-0.52, -0.44) |
| New Mexico | 0.222 | (0.18, 0.26) | 0.078 | (-0.01, 0.17) | -0.144*** | (-0.2, -0.09) |
| New York | 0.458 | (0.41, 0.51) | -0.051 | (-0.18, 0.08) | -0.509*** | (-0.6, -0.42) |
| North Carolina | 0.352 | (0.31, 0.39) | 0.025 | (-0.07, 0.12) | -0.327*** | (-0.39, -0.27) |
| North Dakota | 0.139 | (0.03, 0.25) | 0.07 | (-0.18, 0.32) | -0.07 | (-0.21, 0.07) |
| Ohio | 0.406 | (0.34, 0.47) | 0.05 | (-0.1, 0.2) | -0.357*** | (-0.45, -0.27) |
| Oklahoma | 0.347 | (0.29, 0.4) | -0.002 | (-0.14, 0.13) | -0.348*** | (-0.43, -0.27) |
| Oregon | 0.244 | (0.21, 0.28) | -0.016 | (-0.1, 0.06) | -0.259*** | (-0.31, -0.21) |
| Pennsylvania | 0.405 | (0.37, 0.44) | 0.048 | (-0.04, 0.14) | -0.357*** | (-0.42, -0.3) |
| Rhode Island | 0.371 | (0.29, 0.45) | 0.123 | (-0.06, 0.3) | -0.247*** | (-0.35, -0.15) |
| South Carolina | 0.323 | (0.29, 0.36) | 0.0003 | (-0.09, 0.09) | -0.323*** | (-0.38, -0.27) |
| South Dakota | 0.175 | (0.08, 0.27) | 0.157 | (-0.08, 0.4) | -0.017 | (-0.16, 0.13) |
| Tennessee | 0.281 | (0.23, 0.33) | -0.025 | (-0.16, 0.11) | -0.306*** | (-0.4, -0.22) |
| Texas | 0.316 | (0.29, 0.34) | -0.016 | (-0.1, 0.07) | -0.333*** | (-0.4, -0.27) |
| Utah | 0.312 | (0.27, 0.35) | 0.002 | (-0.09, 0.1) | -0.31*** | (-0.37, -0.25) |
| Vermont | 0.279 | (0.2, 0.36) | -0.124 | (-0.32, 0.07) | -0.403*** | (-0.52, -0.29) |
| Virginia | 0.282 | (0.24, 0.33) | 0.105 | (-0.01, 0.22) | -0.177*** | (-0.25, -0.11) |
| Washington | 0.209 | (0.17, 0.24) | -0.08 | (-0.17, 0.01) | -0.289*** | (-0.35, -0.23) |
| West Virginia | 0.359 | (0.26, 0.46) | 0.014 | (-0.21, 0.24) | -0.345*** | (-0.47, -0.22) |
| Wisconsin | 0.294 | (0.25, 0.34) | -0.013 | (-0.14, 0.11) | -0.307*** | (-0.38, -0.23) |
| Wyoming | 0.177 | (0.13, 0.22) | -0.07 | (-0.18, 0.04) | -0.248*** | (-0.32, -0.18) |

Note: \* $p < 0.1$ ; \*\* $p < 0.05$ ; \*\*\* $p < 0.01$ for corresponding one-sided $p$ -values

**Figure 4:**
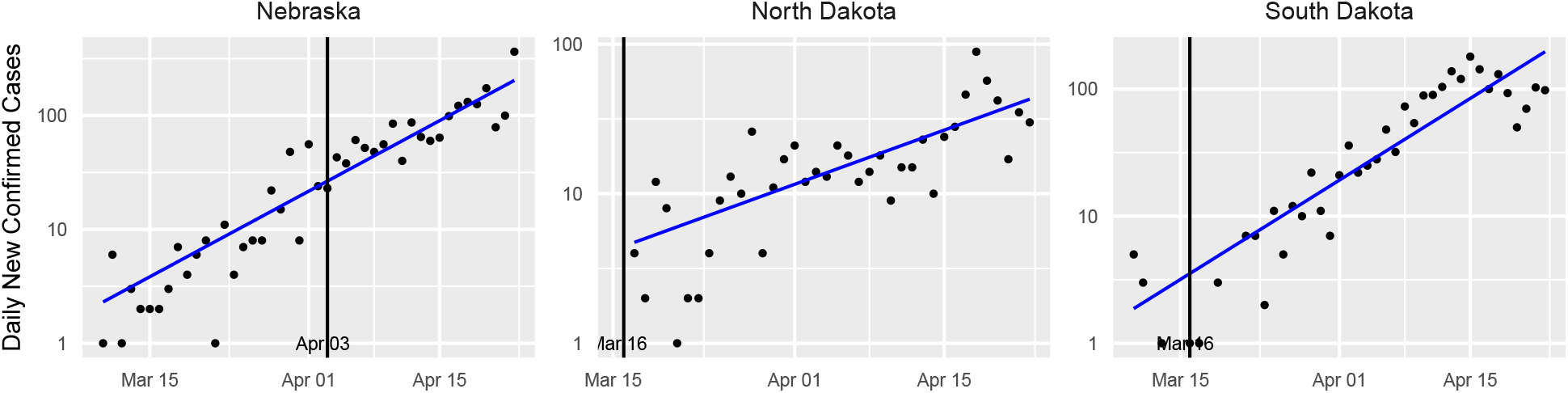
Confirmed case counts for states without a detected changepoint. The vertical black line indicates the intervention date for each state.

**Figure 5:**
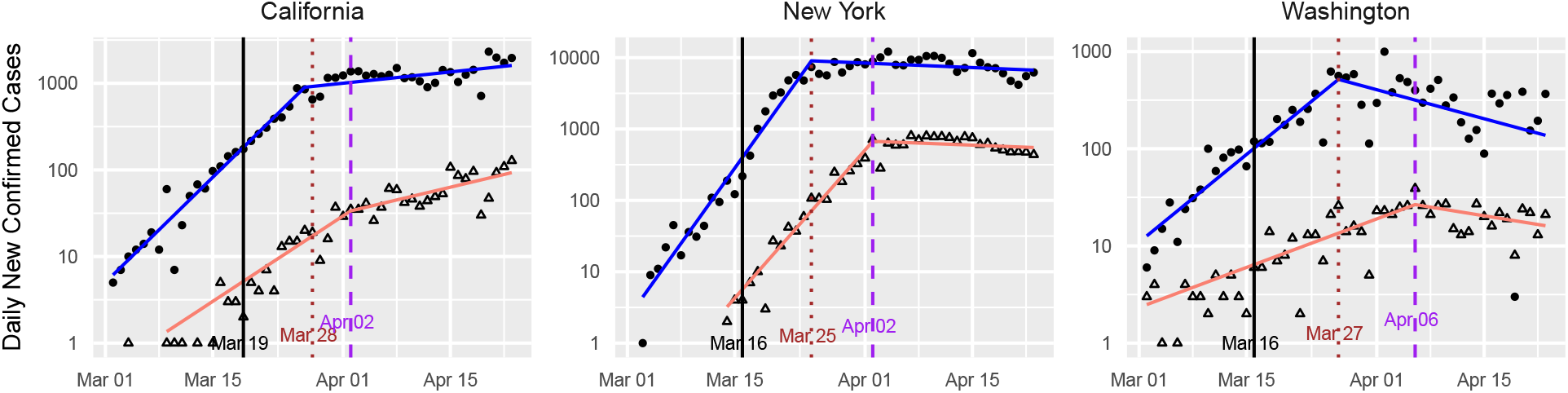
Standard plots for California, New York, and Washington State, augmented with data on confirmed deaths. The vertical black line indicates the intervention date for each state. The sloped lines indicate the fitted values from the learned-delay method applied to the confirmed case data (blue) and the confirmed deaths data (red).

**Figure 6:**
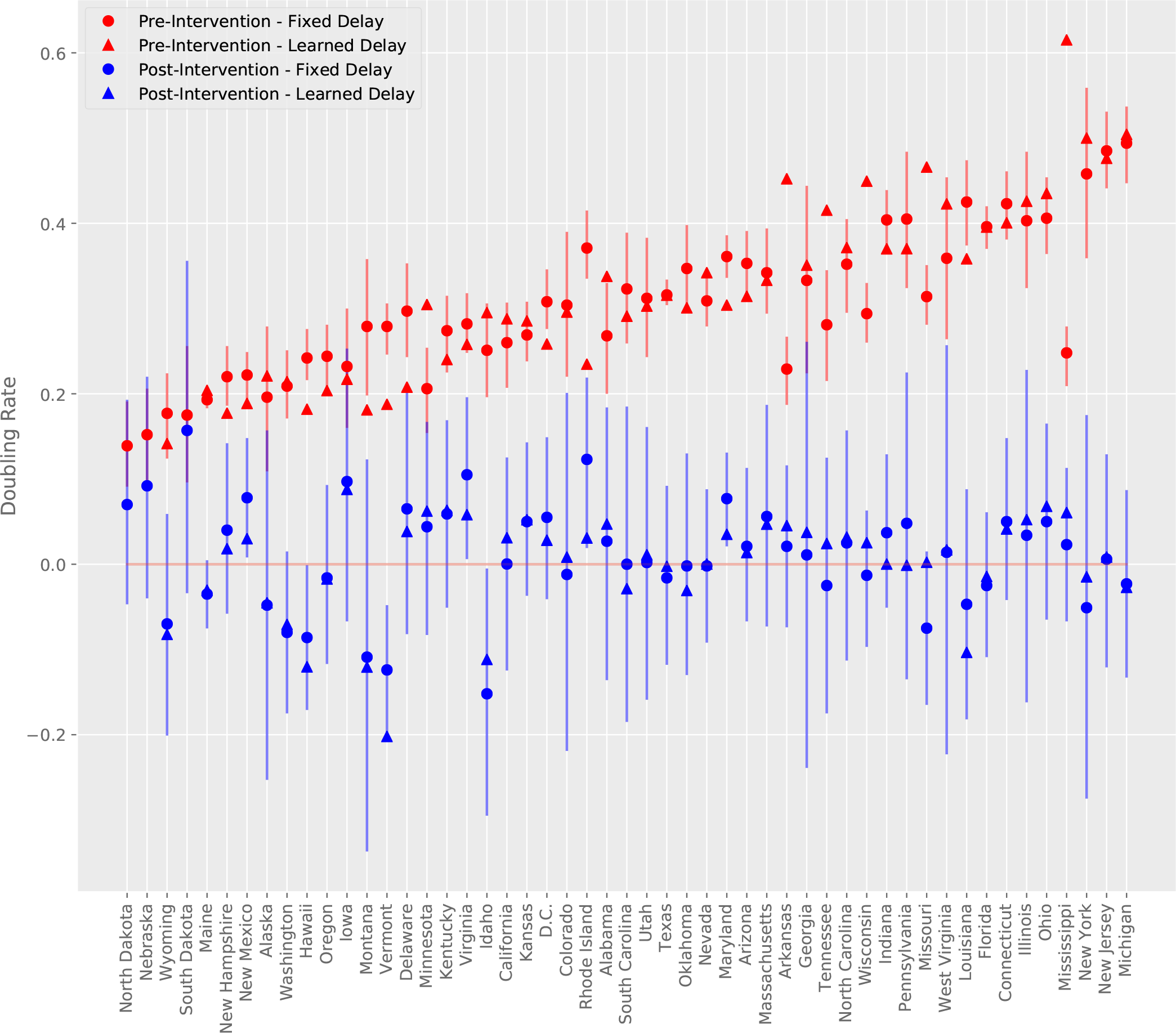
Doubling rate estimates for all states pre-intervention and post-intervention under learned-delay and fixed-delay models. States are in increasing order with respect to the mean of pre-intervention doubling rates under the fixed-delay and learned-delay models. Vertical segments depict approximate 95% confidence intervals for the fixed-delay model estimates.

Table 1 includes estimates of the confirmed case delay, *N*, for those states for which a change-point is detected by the learned-delay model. This column of the table is plotted in Fig. 2, and a histogram is provided in Fig. 3. The median is 11.5 days, which supports the choice used in the fixed-delay model, as noted earlier. the model declines to declare a change-point for three states, whose standard plot is shown Fig. 4. For such states, this model does not find evidence of the efficacy of the social distancing measures imposed. For Nebraska in particular, the intervention occurred quite late (Apr. 3rd), so it is possible that the change is simply not observed in the available data.

Standard plots (showing the fitted learned-delay model) for each state are provided in Figs. 11-14 in Appendix B. As exemplars, we consider three states that were among the first in the US to have confirmed cases, namely New York, California, and Washington. These states have large numbers of confirmed cases, making the data from these states of higher quality. Since the number of deaths is at least an order of magnitude lower than the number of cases, for most states the former is too noisy to be of much use. For these states, however, it provides a useful comparison with the confirmed case counts.

**Figure 7:**
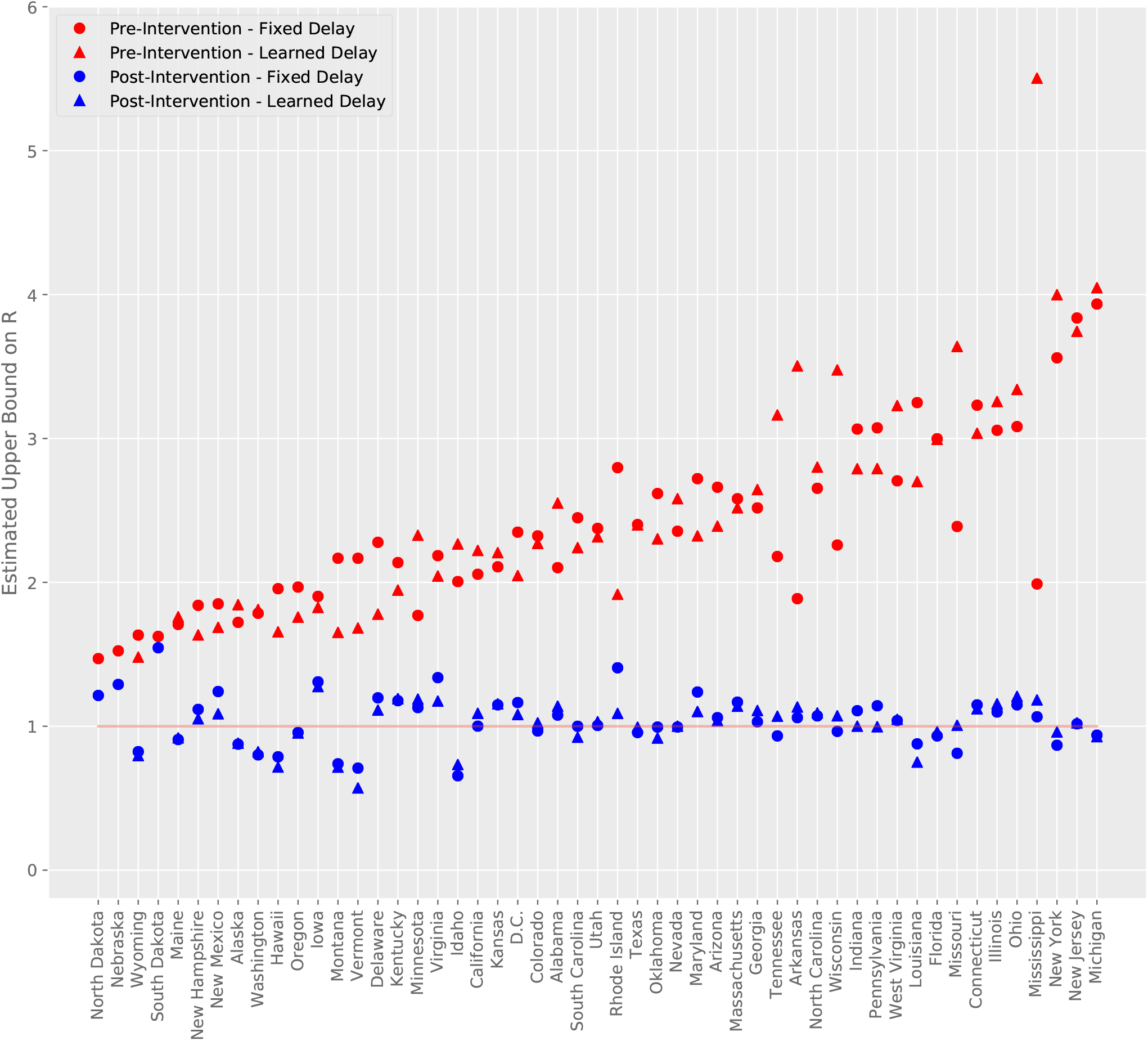
Estimated upper bound on *R* for all states pre-intervention and post-intervention under learned-delay and fixed-delay models. States are in increasing order with respect to the mean of pre-intervention doubling rates under the fixed-delay and learned-delay models.

**Figure 8:**
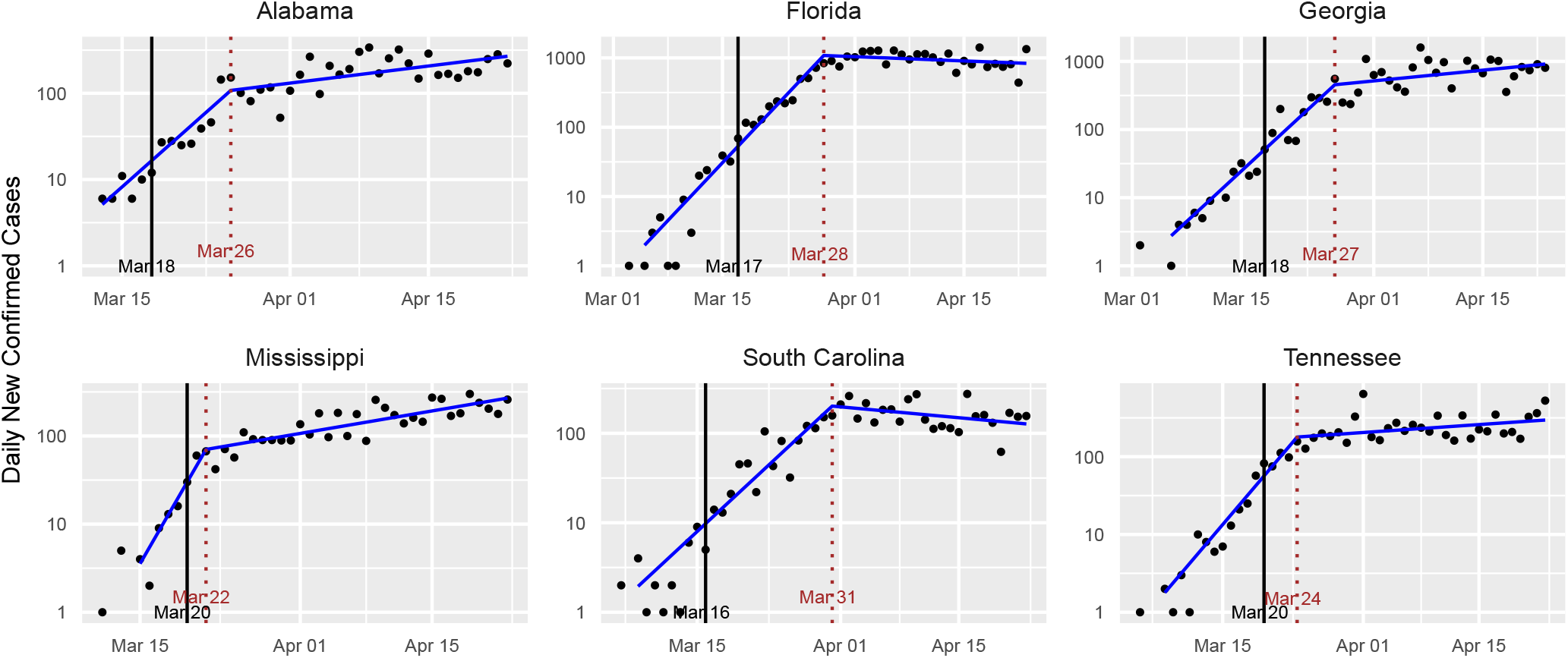
Standard plots for several states proposing an imminent relaxation of social distancing measures as of April 26th.

**Figure 9:**
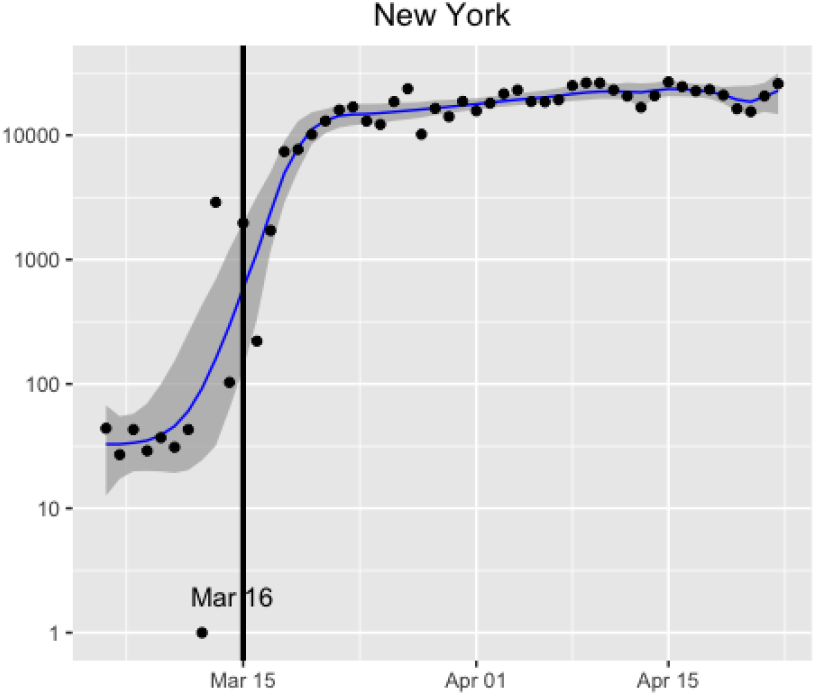
The daily number of tests in New York State. The vertical black line indicates the intervention date. The blue line indicates asmoothed trend estimated by the dynamic shrinkage model ([37]), with corresponding dark gray (credible) bands reflecting variability in the trend estimate.

**Figure 10:**
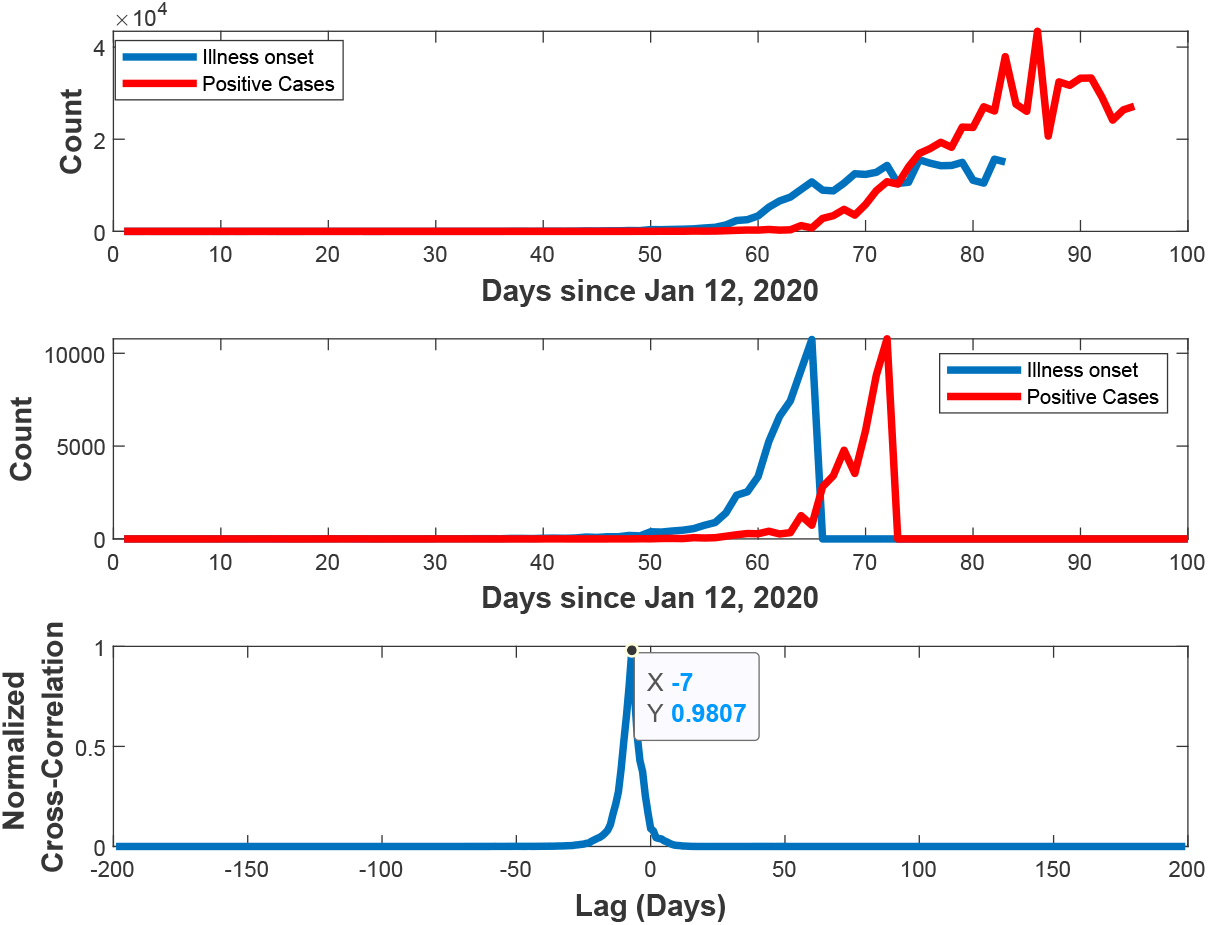
Estimation of confirmed case delay via cross-correlation between illness-onset and positive case confirmation time series.

**Figure 11:**
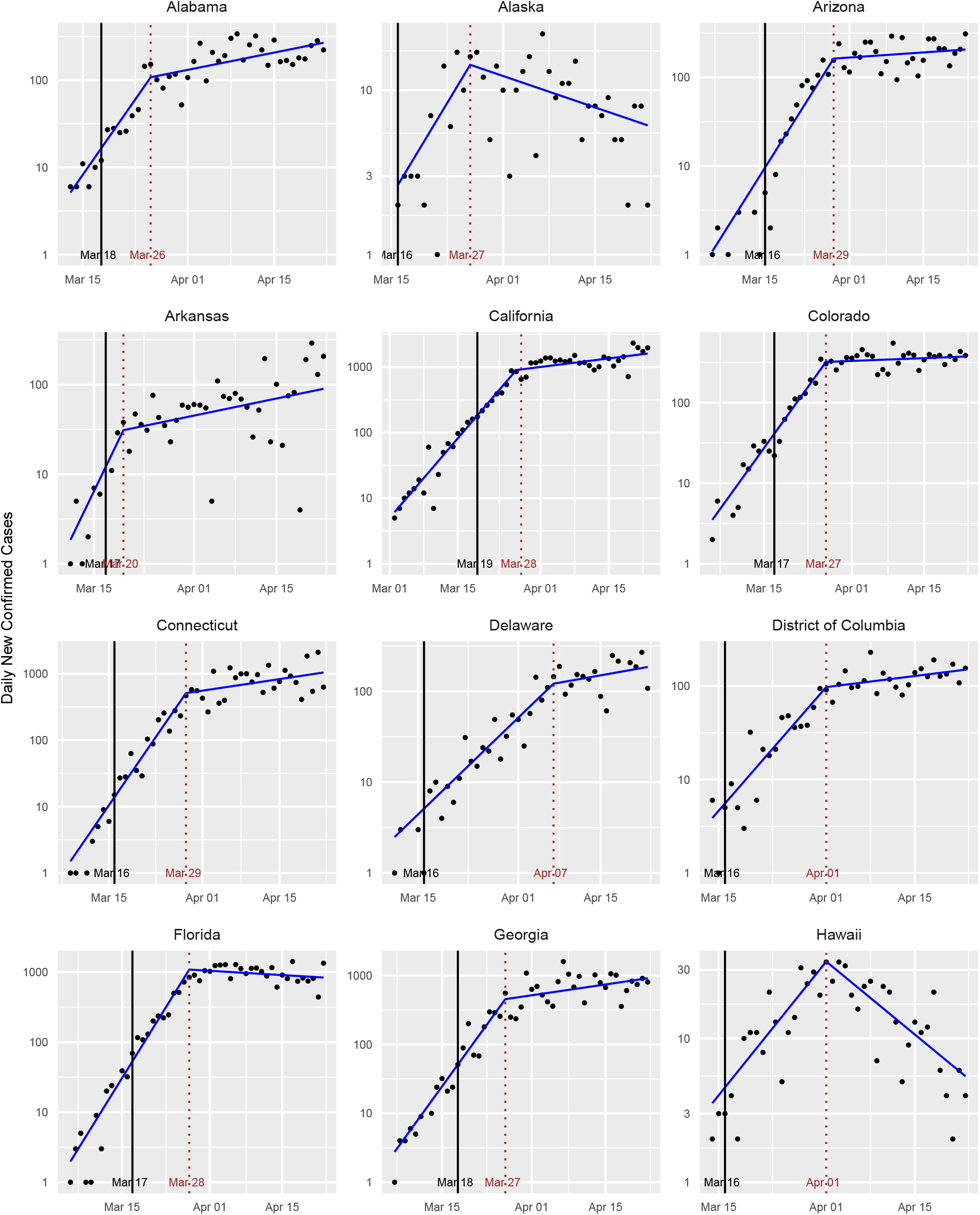
Standard plots for various states.

**Figure 12:**
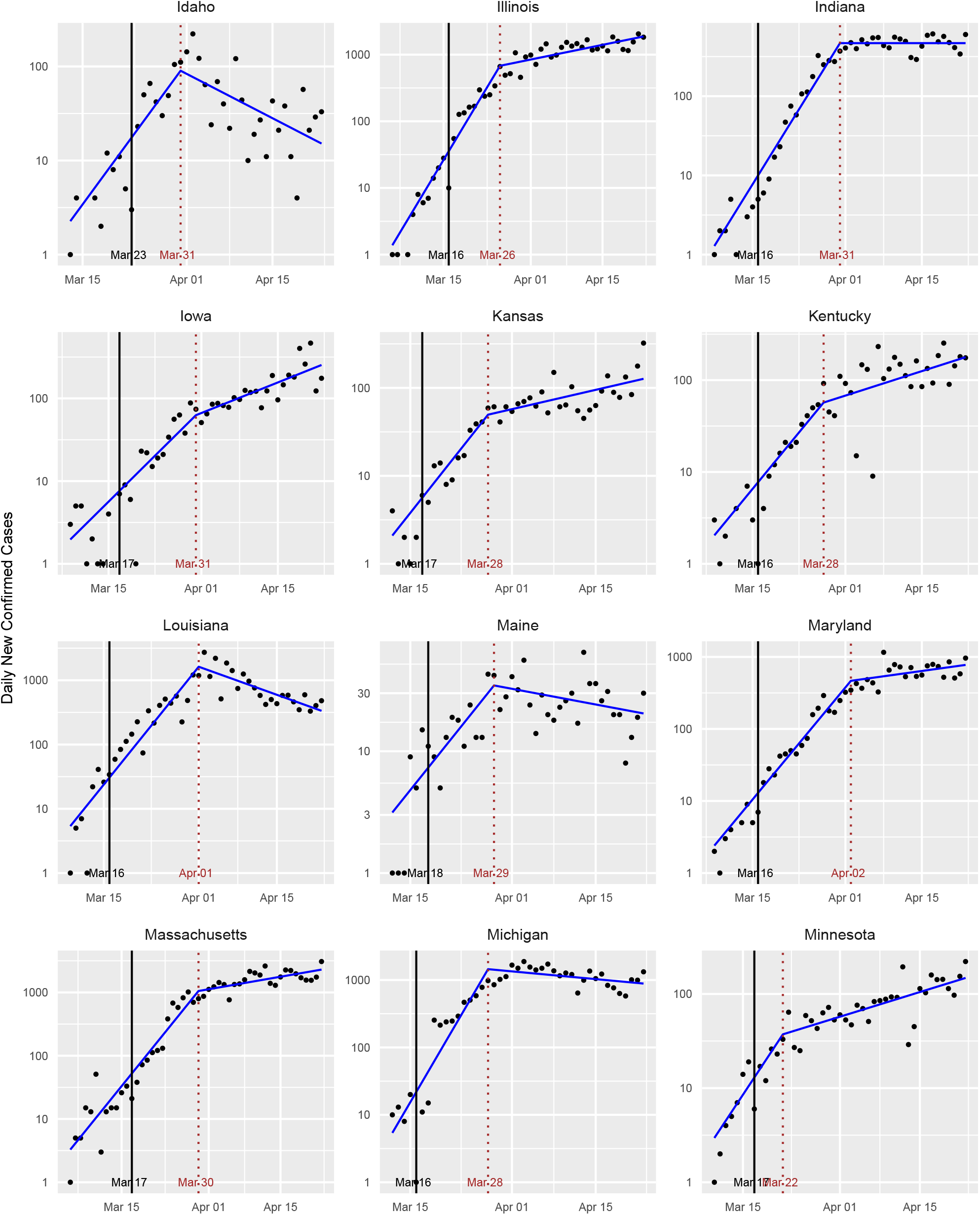
Standard plots for various states.

**Figure 13:**
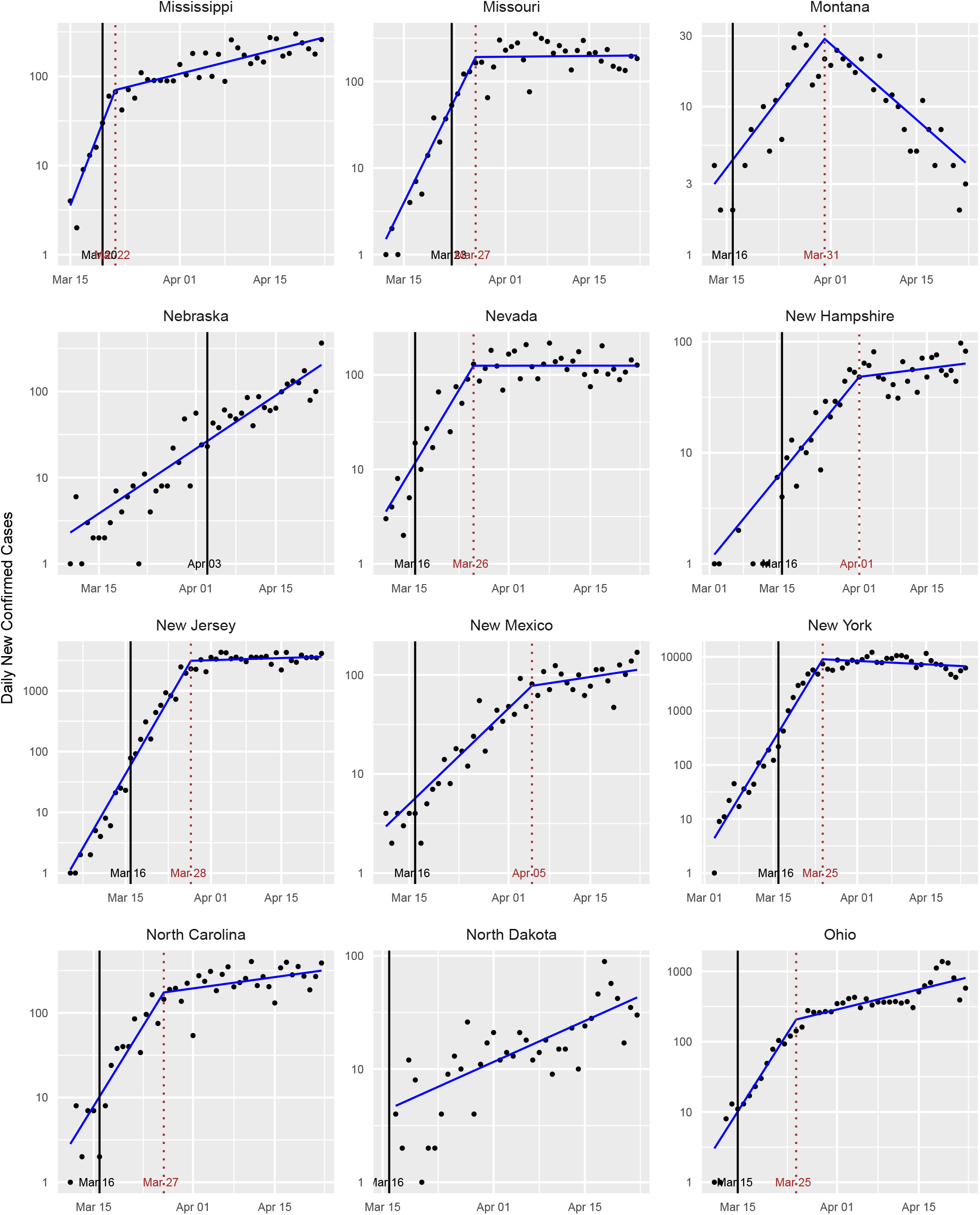
Standard plots for various states.

**Figure 14:**
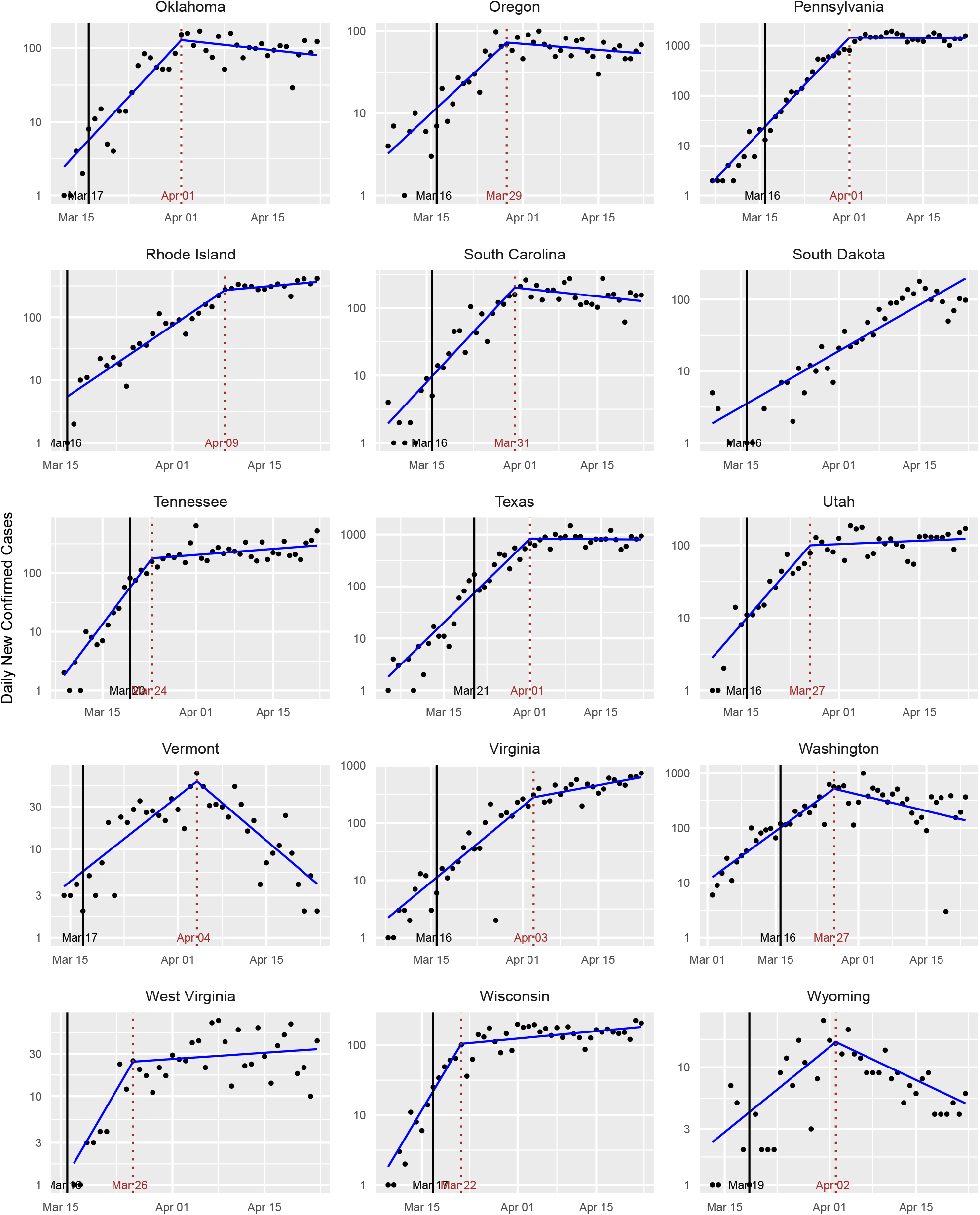
Standard plots for various states.

The results of our analysis for these three states is shown in Figure 5. For all three states, we see two regimes for the growth rate for both the confirmed cases and deaths data. The growth rate begins in asupercritical state indicating exponential growth. The growth rate then decreases substantially, presumably indicating that the intervention is making an impact, albeit with a delayed effect. However, we note that the number of confirmed cases and deaths does not rapidly decline after the change for California and New York, whereas the Washington plot appears to show a decrease in both cases and deaths as a result of the intervention.

To determine the significance level of this finding, we turn to the fixed-delay model, which provides approximate 95% confidence intervals for the doubling rates and their difference (see Table 2). It also provides approximate *p*-values against the null hypothesis that *β*_1_ *≥* 0 and *β*_1_ *≥ β*_0_. In the latter case, we find that the intervention is associated with astatistically-significant decrease in the doubling rate (*p <* 0.01) for all states, even after controlling for false discovery rates using the Benjamini-Hochberg procedure [32], except for Nebraska, South Dakota, and North Dakota. In the former case, when controlling for false discovery, we are not able to conclude that any of the states have achieved a negative *β*_1_; Washington comes the closest with respect to statistical significance.

The point estimates across the ensemble of states also show asubstantial reduction in the doubling rate associated with the intervention, with the post-intervention doubling rate being close to critical. The pre-intervention doubling-rate averaged across all states is 0.302. Post-intervention, it is 0.010, the difference being 0.292. Using Eq. (2), these translate to estimated upper bounds on *R* pre- and post-intervention of 2.310 and 1.028, respectively, or a 55% reduction in contact between contagious and susceptible individuals. It should be emphasized, however, that there is a considerable variation in these values among the states, and the *R* induced from the average doubling rate is distinct from the average of the induced *R* values. Estimated bounds on *R* for each state are provided in Table 3 and Fig. 7.

**Table 3:**
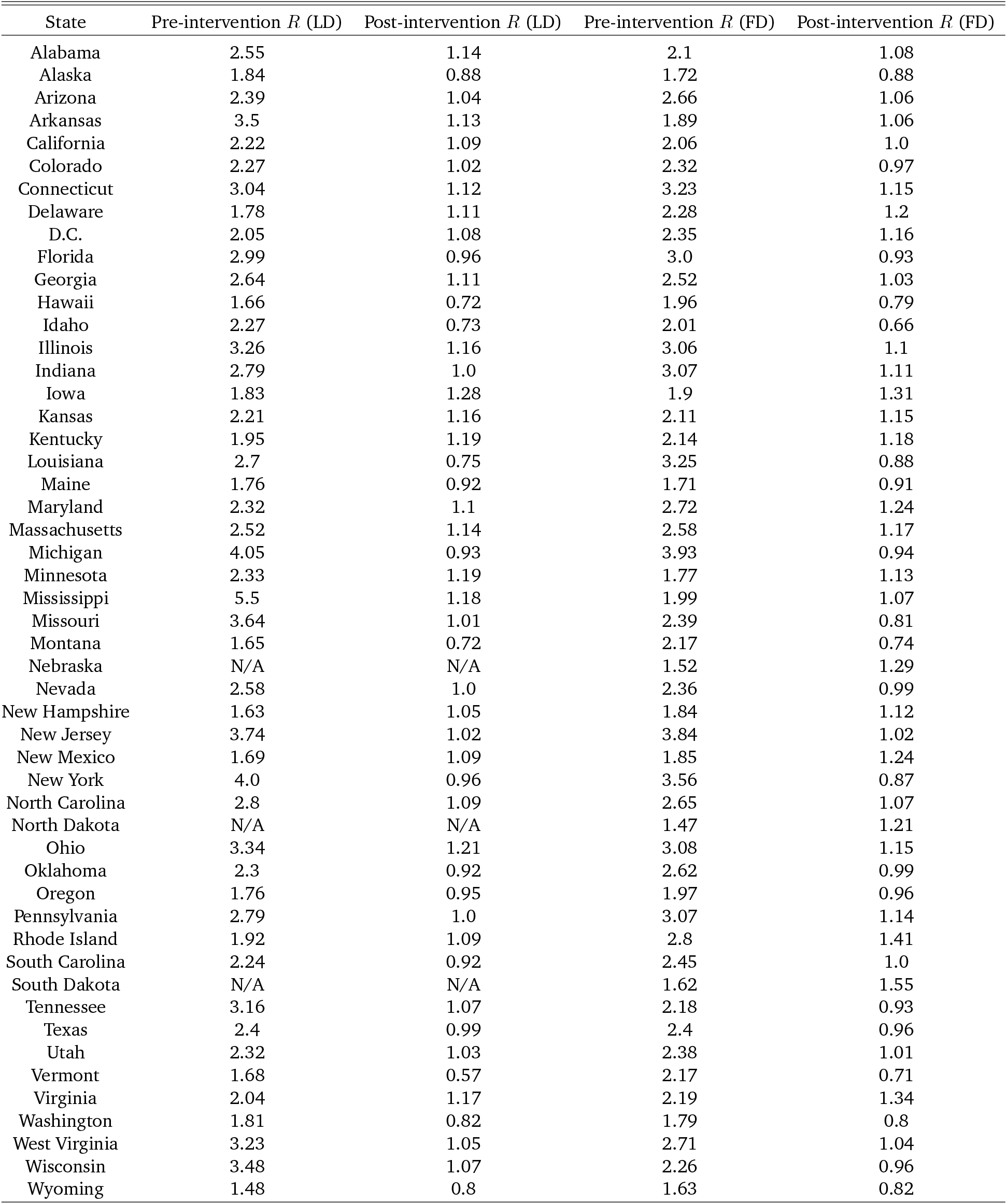
Estimated upper bound on *R* from the Learned-Delay (LD) and Fixed-Delay (FD) models.

**Table 4:** Intervention date by state. The intervention date is defined to be the earlier of the K-12 closure date and the restaurant closure date.

| State | K-12 School Closure | Restaurant Closure | Intervention Date |
| --- | --- | --- | --- |
| Alabama | 2020-03-18 | 2020-03-19 | 2020-03-18 |
| Alaska | 2020-03-16 | 2020-03-18 | 2020-03-16 |
| Arizona | 2020-03-16 | 2020-03-20 | 2020-03-16 |
| Arkansas | 2020-03-17 | 2020-03-20 | 2020-03-17 |
| California | 2020-03-19 | 2020-03-19 | 2020-03-19 |
| Colorado | 2020-03-23 | 2020-03-17 | 2020-03-17 |
| Connecticut | 2020-03-17 | 2020-03-16 | 2020-03-16 |
| Delaware | 2020-03-16 | 2020-03-16 | 2020-03-16 |
| D.C. | 2020-03-16 | 2020-03-16 | 2020-03-16 |
| Florida | 2020-03-17 | 2020-03-20 | 2020-03-17 |
| Georgia | 2020-03-18 | 2020-04-03 | 2020-03-18 |
| Hawaii | 2020-03-16 | 2020-03-17 | 2020-03-16 |
| Idaho | 2020-03-23 | 2020-03-25 | 2020-03-23 |
| Illinois | 2020-03-17 | 2020-03-16 | 2020-03-16 |
| Indiana | 2020-03-19 | 2020-03-16 | 2020-03-16 |
| Iowa | 2020-04-02 | 2020-03-17 | 2020-03-17 |
| Kansas | 2020-03-17 | 2020-03-20 | 2020-03-17 |
| Kentucky | 2020-03-16 | 2020-03-16 | 2020-03-16 |
| Louisiana | 2020-03-16 | 2020-03-17 | 2020-03-16 |
| Maine | 2020-04-02 | 2020-03-18 | 2020-03-18 |
| Maryland | 2020-03-16 | 2020-03-16 | 2020-03-16 |
| Massachusetts | 2020-03-17 | 2020-03-17 | 2020-03-17 |
| Michigan | 2020-03-16 | 2020-03-16 | 2020-03-16 |
| Minnesota | 2020-03-18 | 2020-03-17 | 2020-03-17 |
| Mississippi | 2020-03-20 | 2020-03-24 | 2020-03-20 |
| Missouri | 2020-03-23 | 2020-03-23 | 2020-03-23 |
| Montana | 2020-03-16 | 2020-03-20 | 2020-03-16 |
| Nebraska | 2020-04-03 | 2020-04-03 | 2020-04-03 |
| Nevada | 2020-03-16 | 2020-03-21 | 2020-03-16 |
| New Hampshire | 2020-03-16 | 2020-03-16 | 2020-03-16 |
| New Jersey | 2020-03-18 | 2020-03-16 | 2020-03-16 |
| New Mexico | 2020-03-16 | 2020-03-19 | 2020-03-16 |
| New York | 2020-03-18 | 2020-03-16 | 2020-03-16 |
| North Carolina | 2020-03-16 | 2020-03-17 | 2020-03-16 |
| North Dakota | 2020-03-16 | 2020-03-20 | 2020-03-16 |
| Ohio | 2020-03-16 | 2020-03-15 | 2020-03-15 |
| Oklahoma | 2020-03-17 | 2020-04-01 | 2020-03-17 |
| Oregon | 2020-03-16 | 2020-03-17 | 2020-03-16 |
| Pennsylvania | 2020-03-16 | 2020-03-19 | 2020-03-16 |
| Rhode Island | 2020-03-16 | 2020-03-17 | 2020-03-16 |
| South Carolina | 2020-03-16 | 2020-03-18 | 2020-03-16 |
| South Dakota | 2020-03-16 |  | 2020-03-16 |
| Tennessee | 2020-03-20 | 2020-03-23 | 2020-03-20 |
| Texas | 2020-03-23 | 2020-03-21 | 2020-03-21 |
| Utah | 2020-03-16 | 2020-03-19 | 2020-03-16 |
| Vermont | 2020-03-18 | 2020-03-17 | 2020-03-17 |
| Virginia | 2020-03-16 | 2020-03-25 | 2020-03-16 |
| Washington | 2020-03-17 | 2020-03-16 | 2020-03-16 |
| West Virginia | 2020-03-16 | 2020-03-18 | 2020-03-16 |
| Wisconsin | 2020-03-19 | 2020-03-17 | 2020-03-17 |
| Wyoming | 2020-03-19 | 2020-03-19 | 2020-03-19 |

Thus while this study finds social distancing measures to be effective at reducing the spread of SARS-CoV-2, it does not find conclusive evidence that they have pushed the spread into the subcritical (*β*_1_ *<* 0) regime. Across the ensemble of states, the post-intervention slopes are in fact quite close to zero. The mean slope of the point estimates is 0.010, as noted earlier, with 31 of the states having a post-intervention doubling rate between *−* 0.05 and 0.05, which corresponds to a doubling or halving time exceeding twenty days. This indicates that the pandemic across many regions plateaued, rather than contracted, post-intervention. In those locations, the spread is now behaving akin to a critical branching process [12], with new cases replacing recoveries and deaths. This plateau should be contrasted with the symmetrical apex that is presumed in some predictive models [33].

## 5 Policy Implications

Due to the economic and social costs of social distancing, there is interest in the question of when states should relax such measures. Some states have already begun this process with Georgia allowing certain businesses, such as gyms and barbershops, to reopen [34]. In fact, Alabama, Florida, Georgia, Mississippi, South Carolina and Tennessee announced a coordinated attempt to reopen [35, 36]. Standard plots for these states are shown in Fig. 8.

Since asystematic relaxation of social distancing will presumably increase the doubling rate, from a public health perspective it is advisable to relax such measures only when there is evidence that the spread has become subcritical. Among negative doubling rates, those that are farther from zero (i.e., are larger in absolute value) allow for more relaxation of social distancing measures without the spread returning to supercriticality.

We see that the curves for these states appear similar to those for the early states in Fig. 5. In particular, we see asubstantial change in the growth rate occurring some days after the intervention, but the spread does not appear to have become subcritical, with the exception of South Carolina and possibly Florida. This study finds that other states, such as Idaho, Vermont, Montana, Washington, and Hawaii have astronger basis for relaxing social distancing at this time.

The lack of clear subcriticality among the post-intervention doubling rates suggests that existing social distancing measures will need to remain in place for some time. On the other hand, it is possible that asubset of the existing social distancing measures are responsible for the observed reductions in the doubling rate. If this is the case, then the remaining measures could be relaxed with no harmful effect. Also, any additional measures taken, such as contract tracing, will tend to reduce the doubling rate further. Finally, in the absence of a change in interventions, the doubling rate will tend to decrease over time as the population of susceptible individuals decreases.

## 6 Limitations of the Analysis

Our analysis relies on the number of confirmed cases of COVID-19 reported by federal, state, territorial, and local authorities. Beyond the delay between infection and confirmation, which was discussed earlier, this statistic is confounded by variability in the number of tests performed. New York State, for instance, saw a rapid increase in the number of daily tests followed by a plateauing around March 19th, as shown in Fig. 9. This approximately coincides with the detected change-point in the number of confirmed cases by the learned-delay method. It is possible that for some states, the abrupt change that we detect in confirmed cases does not reflect an actual change in the spread of disease but only a change in detection capabilities.

Controlling for this effect is challenging because the data on the number of tests is subject to additional types of errors. States vary in whether their test counts report the number of distinct people tested or the number of tests performed, and whether they represent the number of tests taken or completed. Data on the number of tests is also not available for all of the states for the time interval considered here, as some states initially reported only confirmed cases.

One could potentially rely on the number of deaths attributed to COVID-19 instead of the number of confirmed cases. This approach is subject to three complications, however. First, the mean time between infection and death is presumably longer than the time between infection and confirmed diagnosis. Second, the number of deaths is a noisier signal because the number of deaths is lower than the number of confirmed cases, often by an order of magnitude, as noted earlier. Finally, some states have not been performing COVID-19 tests post-mortem. In this case, any reported COVID-19 death would have been already reported as a confirmed case and would not reveal new information about the number of infections.

Mitigating this concern, we note that for New York, the number of tests plateaued slightly before the number of cases did. We also note that data on deaths and hospitalizations, while lower in quality than the number of confirmed cases, are broadly consistent with the findings based on the confirmed case count alone. Notably, we do not see pronounced increases or decreases in deaths or hospitalizations post-intervention for states in which the estimated doubling rate post-intervention is near zero. We also do not see asaturation in the fraction of positive tests, which would indicate an uncaptured exponential increase in the number of actual COVID-19 patients. Finally, several states have continued to see supercritical growth post-intervention.

Another limitation of our analysis is that it treats states separately and thereby ignores the influence that neighboring states have on one another. Nor does it capitalize on the potential similarity among the model parameters for similar states. We also assume an unchanging reservoir of susceptible individuals; an SIR-type model (e.g., [38]) is appropriate when the fraction of the population that is exposed to the virus varies over the period under study. Finally, our analysis assumes asingle changepoint when the first large-scale social distancing measures are imposed. It cannot distinguish among the relative benefits of different interventions imposed around the same time. It also does not account for anticipatory behavioral changes that could have preceded the formal imposition of interventions.

Subsequent analyses may require additional changepoints as states relax, and possibly reimpose, social distancing measures. The confirmed case delay could also vary over time as a result of changing testing protocols.

## Data Availability

All of the data used in this study is publicly available via the provided links.

https://github.com/nytimes/covid-19-data

https://www.cdc.gov/coronavirus/2019-ncov/cases-updates/previouscases.html

https://covidtracking.com

## 7 Acknowledgment

This research was supported in part by the Greater Data Science Cooperative Institute through National Science Foundation grants 1934985 and 1934962 and in part by National Institutes of Health grant DP5OD021338.

## 9 Data availability

The data used are publicly available from https://github.com/nytimes/covid-19-data, https://www.cdc.gov/coronavirus/2019-ncov/cases-updates/previouscases.html, and https://covidtracking.com.

## Appendices

### A An Independent Estimate of the Confirmed Case Delay

The quantity *N* is the sum of the mean incubation time and the mean time between symptom onset and confirmed diagnosis. Various works have estimated the mean incubation time to be 5 days [39, 40, 41]. We focus here on estimating the time between symptom onset and confirmed diagnosis.

We estimate this quantity using data from the CDC [24]. The CDC reports COVID-19 cases in the United States by date of illness onset from January 12, 2020, to April 15, 2020, as well as by date of confirmed diagnosis (Fig. 10, top). The CDC warns that estimates are not accurate after April 5, 2020 due to difficulty in accurately determining illness onset after community spread began.

We truncate the time-series so that both end when their respective cumulative counts reach 10,000 (Fig. 10, middle). The lag between illness onset and case confirmation is determined using a normalized cross-correlation operation (Fig. 10, bottom). The lag is estimated to be 7 days. When added to the incubation time, this results in a total time of 12 days from exposure to confirmed test, which essentially coincides with the estimate of 11.5 days obtained in the body of the paper. An earlier work estimated the mean time from symptom onset to confirmed test to be 4.8 days [42], which translates to a point estimate of *N* of about 10 days.

### B Parameter Estimates and Standard Plots for All States

## Footnotes

1 For convenience, we shall refer to all of these as “states.”

2 If one assumes equality in (2) and adopts the choice *µ* = 4 days, then one arrives at the following rule-of-thumb for estimating *R* for COVID-19 in a particular locale: divide the number of new cases reported today by the number of new cases reported four days ago. Another rule, which is more impervious to errors due to differences in reporting over different days of the week, is to take the square root of the ratio of the number of cases reported today to the number of cases reported one week ago. The former rule is applicable if *R* has been constant for at least the *confirmed case delay* (which is defined shortly) plus 4 days. The latter rule is applicable if *R* has been constant for at least the confirmed case delay plus 7 days.

